# Seroprevalence of SARS-CoV-2 in Niger State: A Pilot Cross Sectional Study

**DOI:** 10.1101/2020.08.04.20168112

**Authors:** Hussaini Majiya, Mohammed Aliyu-Paiko, Vincent T Balogu, Dickson A Musa, Ibrahim M Salihu, Abdullahi A Kawu, Yakubu I Bashir, Aishat A Sani, Baba John, Amina T Muhammad, Fatima L Jibril, Ezekiel Bala, Nuhu G Obaje, Yahaya B. Aliyu, Ramatu G Muhammad, Hadiza Mohammed, Usman N Gimba, Abduljelili Uthman, Hadiza M Liman, Alfa A Sule, Kolo J Joseph, Mohammed M Makusidi, Mohammed D Isah, Ibrahim Abdullahi, Umar Ndagi, Bala Waziri, Chindo I Bisallah, Naomi J Dadi-Mamud, Kolo Ibrahim, Abu Kasim Adamu

**Author notes:** Address correspondence to Hussaini Majiya, Department of Microbiology, Ibrahim Badamasi Babangida University, Lapai, KM3 Lapai-Minna Road, P.M.B 11, Lapai, Nigeria.

## Abstract

**Background:** Coronavirus Disease 2019 (COVID-19) Pandemic caused by SARS-CoV-2 is ongoing causing human and socioeconomic losses.

**Objective:** To know how far the virus has spread in Niger State, Nigeria, a pilot study was carried out to determine the SARS-CoV-2 seroprevalence, patterns, dynamics, and risk factors in the state.

**Methods:** A cross sectional study design and Clustered-Stratified-Random sampling strategy were used to select 185 test participants across the state. SARS-CoV-2 IgG and IgM Rapid Test Kits (Colloidal gold immunochromatography lateral flow system) were used to determine the presence or absence of the antibodies to the virus in the blood of sampled participants across Niger State as from 26th June 2020 to 30th June 2020. The test kits were validated using the blood samples of some of the Nigeria Center for Disease Control (NCDC) confirmed positive and negative COVID-19 cases in the State. SARS-CoV-2 IgG and IgM Test results were entered into the EPIINFO questionnaire administered simultaneously with each test. EPIINFO was then used for to calculate arithmetic mean and percentage, odd ratio, chi-square, and regression at 95% Confidence Interval of the data generated.

**Results:** The seroprevalence of SARS-CoV-2 in Niger State was found to be 25.41% and 2.16% for the positive IgG and IgM respectively. Seroprevalence among age groups, gender and by occupation varied widely. COVID-19 asymptomatic rate in the state was found to be 46.81%. The risk analyses showed that the chances of infection are almost the same for both urban and rural dwellers in the state. However, health care workers, those that experienced flu-like symptoms and those that have had contact with person (s) that travelled out of Nigeria in the last six (6) months (February -June 2020) are twice (2 times) at risk of being infected with the virus. More than half (54.59%) of the participants in this study did not practice social distancing at any time since the pandemic started. Discussions about knowledge, practice and attitude of the participants are included.

**Conclusion:** The observed Niger State SARS-CoV-2 seroprevalence and infection patterns means that the virus is widely spread, far more SARS CoV-2 infections occurred than the reported cases and high asymptomatic COVID-19 across the state.

## Introduction

COVID-19 Pandemic is caused by a novel coronavirus-SARS-CoV-2 that is believed to have crossed from bats to humans for the first time [1-3]. COVID-19 is an infectious disease of respiratory system of humans and animals and the virus can be transmitted through facial openings which include mouth, nostrils and maybe eyes [2-4].

The first case of COVID-19 in Niger State, Nigeria, was announced by the Nigeria Center for Disease Control (NCDC) on 10^th^ April 2020; this was after about six (6) weeks when the first confirmed case (index case) of COVID-19 in Nigeria was announced on 27^th^ February 2020 when a foreigner in Lagos tested positive for SARS-CoV-2. Since then, many cases have been confirmed for the state and it is still increasing [5]. As part of the measures to curtail the spread of SARS CoV-2, strict lockdown (restriction of people to their homes except for essential needs-medicine and food) was enforced in the state as from 25^th^ March 2020 to 9^th^ June 2020. However, full compliance to the strict lockdown by the citizens of the State may not have been achieved or possible, due socio-economic and cultural reasons, disbeliefs, and conspiracy theories. Many people would have to go out on daily basis to work to cater for their families and markets are usually open spaces bustling with large crowds of people. Many people did not believe in the existence of COVID-19 and especially about the highly contagious nature of the disease. There is also no efficient and robust housing and biometric data management systems where everyone is accounted for, especially for the purposes of employment, health, security and social welfare. If these are available, foods and other goods purchased online can be sent to houses with ease. Also, utilities are in most cases not provided or grossly inadequately supplied. It is difficult for people to stay back at home and comply with the strict lockdown in such situations. After the ease of lockdown, there has been enhanced enforcement of the compulsory use of face mask in public places and adherence to physical distancing in the state [5].

Niger State is one of the thirty-six (36) states in Nigeria and has Minna as its capital. It has twenty-five (25) Local Government Areas which are fairly distributed among the three (3) geopolitical zones of the state in terms of land mass and populations. In terms of land mass, Niger state is the largest state (76,363 km^2^) in Nigeria and 18^th^ (5,556,247 people) in Nigeria in terms of population [6,7]. However, as of 21^st^ December 2020, Niger State is ranked 28^th^ among the list of states for COVID-19 cases reported in Nigeria. The total reported COVID-19 cases in the state as of 21^st^ December 2020 is 381 with 12 deaths while for Nigeria (with a population of about 206,630,269) is 79, 789 COVID-19 cases and 1,231 deaths [5-8]. It is generally believed that the reported COVID-19 cases in the state and Nigeria are very far below the actual SARS CoV-2 infections in the populations. This may be due to Polymerase Chain Reaction (PCR) based SARS CoV-2 test limitations in many states of Nigeria and unknown proportion of mild or asymptomatic COVI-19 cases that may not be diagnosed and not reported. Presence and detection of antibodies to SARS-CoV-2 in the blood of sampled participants will likely indicates they were infected at some point since the start of the pandemic. Therefore, serologic assays can be used to determine population-based estimates of SARS CoV-2 infection, including those who had mild or asymptomatic infection or who were never tested despite symptoms [9-12].

Coronavirus Disease 2019 (COVID-19), like most of infectious diseases, the isolation of the aetiologic agent-SARS CoV-2 through tissue/cell plate culture technique would have been the gold standard method for the diagnostic test. However, plate culturing is usually laborious, time consuming, complex and costly and therefore impossible to use especially for epidemiological studies where large samples may be involved. Also, even though Reverse Transcriptase-Polymerase Chain Reaction (RT-PCR) have been predominantly used to test for the agent of COVID-19 world over including Nigeria, it is laborious, time consuming, costly and complex [13-15].

Infection by many pathogens including viruses do elicit production of antibodies in humans and animals even if no symptoms manifested. The detection of the antibodies in the whole blood/serum/plasma of humans and animals have been used as preliminary diagnoses of infectious diseases [9-17].Also because of the relative ease of use and simplicity of the antigen-antibodies test kits compared to cell/tissue culture and PCR, they are mostly used in the epidemiological studies to determine infectious diseases prevalence, patterns, dynamics, and risk factors [13-17]. Antigen-Antibody based kits unlike other methods can detect previous exposure to the infectious agents [9-17]-this information is very important especially in COVID-19 with assumed high rate of asymptomatic cases in order to see how far the virus has spread and infection patterns, effectiveness of social distancing measures enforced. This pilot study was aimed at determining the SARS CoV-2 seroprevalence, patterns and dynamics and risk factors for contracting COVID-19 in Niger State, Nigeria. It was also aimed at assessing the knowledge, attitude, and practice of people with regards to the COVID-19 and its control measures in the State.

## Methods

### Study Design and Population

A cross sectional study design and clustered-stratified-random sampling strategy were used. The study area was Niger State and its residents were the study population (Figure 1). Niger State (Figure 1) is one of the federating geopolitical states in Nigeria. It has Minna as its capital. Other major towns are Bida, Kontagora, Suleja, New Bussa, Mokwa, Lapai, and Agaie. The three (3) geopolitical zones (Zone A, Zone B and Zone C) in the state were covered fairly (Figure 1). Places of residents (classified as urban and rural), gender, occupation (classified as health care worker and non-health care worker), age group/range, were the stratifications that were applied in those places chosen in each of the zones (Figure 1). The selected sampling points (Appendix 1) were socio-economic places such as hospitals and primary health care centres, motor parks, markets, village/community heads’ households, Sawmills, schools, etc. Considering the stratifications, people in the selected sampling points were randomly approached and recruited to participated in the study. OpenEpi Toolkit was used to calculate the minimum sample size for the study. Since this is a pilot study and SARS CoV-2 prevalence is not known in Niger State that has a population of about 5 million people, we assumed that, overall, the SARS CoV-2 prevalence would be about 50%, with 95% confidence in the estimate, absolute precision of 10% and design effect (for random-cluster surveys) of 1.0, therefor, the minimum required sample size was 97 participants. A total of 185 participants were enrolled in this study. Among individuals approached during the recruitment for this study, average acceptance/participation rate was 87% (185/212). As from 26^th^ June 2020 to 30^th^ June 2020 and with full consent to participate in the study, samples were taken randomly from 185 participants (almost equally distributed among the 3 geopolitical zones) for SARS CoV-2 IgG and IgM Rapid Tests and questionnaire (created by EPIINFO 7.2.2.6) administered simultaneously.

**Figure 1:**
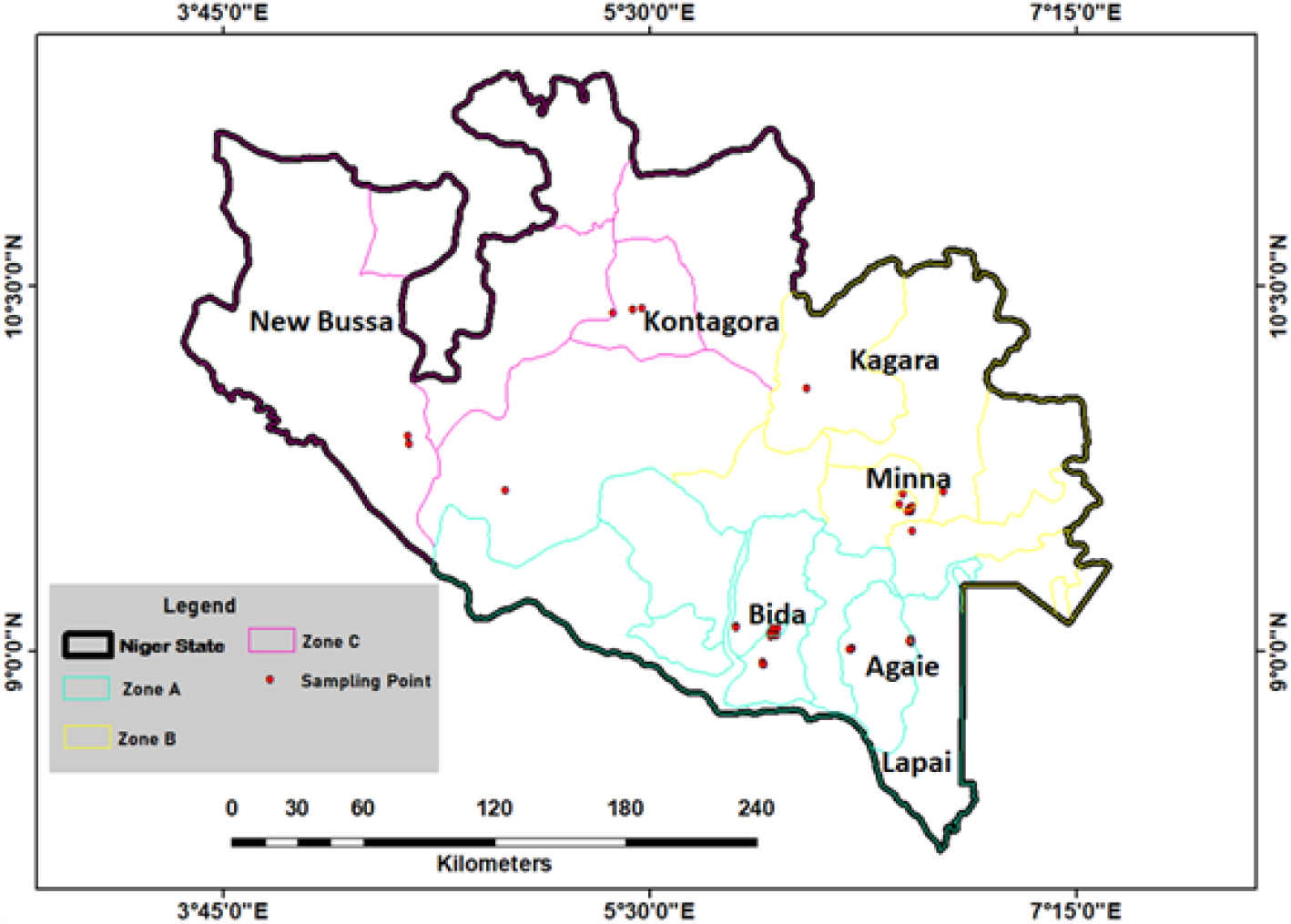
Map of Niger State showing the sites where samples were taken, tests carried out and questionnaire administered in the three (3) geopolitical zones (Zone A, Zone B and Zone C) of the state for the pilot SARS-CoV-2 seroprevalence study carried out as from 26^th^ – 30^th^ June 2020 in Niger State, Nigeria. Exact names, latitudes and longitudes of the sampling points can be found in Appendix 1.

### Ethical Approval and Consent

Ethical approval (STA/495/Vol/152) for this study was given by the Research Ethics Committee of the Niger Sate Ministry of Health. Consents were also sought from each of the participants prior to tests and questionnaire administration, and only those individuals that have given full consent were used in the study. Parents/Guardians were responsible for the consents of their wards that participated in the study and were under 18 of age.

### Specimen Type and SARS CoV-2 IgG and IgM Rapid Test

SARS CoV-2 IgG/IgM Rapid Tests were carried out using the whole blood of the participants. It is a qualitative membrane-based immunoassay for the detection of COVID 19 antibodies in whole blood. The tests were carried out and interpreted according to the kits manufacturer’s instruction. The test result of each participant was recorded and entered into the EPIINFO questionnaire administered for that particular participant. The SARS CoV-2 IgG and IgM Rapid Test Kits were validated with the blood samples of those individuals that were confirmed by the NCDC through PCR as positive and or negative for COVID-19 in Niger State. All the ten (10) confirmed NCDC positive cases sampled tested positive for the IgG for SARS-CoV-2 while all the five (5) confirmed NCDC negative individuals (that have never tested positive before) tested negative for the IgG and IgM for the SARS-CoV-2. This means that 100% sensitivity and specificity were observed for the test kits used in the study.

### EPIINFO Questionnaire and Statistics

In order to be able to determine the COVID-19 prevalence, patterns and dynamics and risk factors for contracting the disease in Niger State, a questionnaire (Appendix 2) was designed and created using EPIINFO 7.2.2.6. The questionnaire was designed to ask some questions with categorical responses (Yes or No) and also to accommodate the test results of the participants; this was to be able to run 2×2 table statistics including calculating SARS CoV-2 infection odd ratios and linear regression (multivariate analysis) for many scenarios. SARS CoV-2 IgG and IgM Test results were entered into the EPIINFO questionnaire administered simultaneously with each test for a participant. EPIINFO was used to calculate arithmetic mean and percentage, odd ratio, chi-square and regression at 95% Confidence Interval of the data generated. Demographics, SARS CoV-2 prevalence and COVID-19 Asymptomatic rate, knowledge, attitude, and practice of the participants were expressed in percentages. For risk of contracting SARS CoV-2 analyses (odd ratio and chi-square), the SARS CoV-2 IgG status (prevalence) was the dependent variable while the demographics and risks were the independent variables.

## Results

### Demographics, Knowledge, Attitude, Practice, Travel History and Flu-like Symptoms of the Participants

The Demographic characteristics, knowledge, attitude, practice, travel history and flu-like symptoms of the participants in this study are shown in Table 1. More than half- 101(54.59%) of the participants in this study did not practice social distancing (Table 1) at any time since the pandemic started (January 2020 to June 2020) even as the lockdown was once enforced in the state. Majority- 114 (61.62%) of the participants practiced hand and face hygiene (Table 1). Vast majority-181 (97.84%) of the participants did not travel out of Nigeria since the beginning of the year 2020 when the pandemic started (Table 1). Only very few- 4(2.16%) of the participants did travel out of Nigeria in the last 6 months (January 2020 to June 2020) and have returned (Table 1). However, 24 (12.97%) of the participants have had contact with someone that travelled out of the country in the last 6 months (January 2020 to June 2020) (Table 1). Majority-113 (61.08%) of the participants did not experience any flu-like symptoms since when the pandemic started (January 2020 to June 2020) (Table 1). Only 72 (38.92%) of the participants experienced flu-like symptoms (January 2020 to June 2020) (Table 1).

**Table 1:** Demographics, knowledge, attitude, practice, travel history and flu-like symptoms of the participants in a pilot SARS-CoV-2 seroprevalence study carried out as from 26^th^ – 30^th^ June 2020 in Niger State, Nigeria.

| Variable | Category | Participants n (%) | Exact 95% LCL (n%) | Exact 95% UCL (n%) |
| --- | --- | --- | --- | --- |
| **Age (Years) (n=185)** |  |  |  |  |
|  | ≤ 5 | 15 (8.11) | 4.61 | 13.02 |
|  | 6-17 | 26 (14.05) | 9.39 | 19.91 |
|  | 18-29 | 34 (18.38) | 13.08 | 24.72 |
|  | 30-41 | 45 (24.32) | 18.33 | 31.16 |
|  | 42-53 | 37 (20.00) | 14.49 | 26.50 |
|  | 54-65 | 20 (10.81) | 6.73 | 16.20 |
|  | ≥66 | 8 (4.32) | 1.89 | 8.34 |
| **Gender (n=185)** |  |  |  |  |
|  | Male | 103 (55.68) | 48.21 | 62.96 |
|  | Female | 82 (44.32) | 37.04 | 51.79 |
| **Resident (n=185)** |  |  |  |  |
|  | Urban | 115 (62.16) | 54.75 | 69.17 |
|  | Rural | 70 (37.84) | 30.83 | 45.25 |
| **Occupation (n=185)** |  |  |  |  |
|  | Health care workers | 43 (23.24) | 17.36 | 30.00 |
|  | Non-Health care workers | 142 (76.76) | 70.00 | 82.64 |
| **Health Care workers: Gender (n=43)** |  |  |  |  |
|  | Male | 22 (51.16) | 35.46 | 66.69 |
|  | Female | 21(48.84) | 33.31 | 64.54 |
| **Knowledge (n=185)** |  |  |  |  |
|  | Aware of COVID-19 | 151 (81.62) | 75.28 | 86.92 |
|  | Not Aware of COVID-19 | 34 (18.38) | 13.08 | 24.72 |
| **Belief (n=185)** |  |  |  |  |
|  | COVID-19 is in Niger State | 109 (58.92) | 51.46 | 66.08 |
|  | COVID-19 is not in Niger State | 76 (41.08) | 33.92 | 48.54 |
| **\*Behavior (A) (n=185)** |  |  |  |  |
|  | Hand and Face Hygiene | 114 (61.62) | 54.20 | 68.66 |
|  | No Hand and Face Hygiene | 71 (38.38) | 31.34 | 45.80 |
| **\*Behavior (B)**<br>**(n=185)** |  |  |  |  |
|  | Social Distancing | 84 (45.41) | 38.09 | 52.87 |
|  | No Social Distancing | 101 (54.59) | 47.13 | 61.91 |
| **\*Travel History**<br>**(n=185)** |  |  |  |  |
|  | Traveled Overseas | 4 (2.16) | 0.59 | 5.44 |
|  | Did not travel overseas | 181 (97.84) | 94.56 | 99.41 |
| **\*Contact History (n=185)** |  |  |  |  |
|  | Contact with overseas returnee | 24 (12.97) | 8.49 | 18.69 |
|  | No contact with overseas returnee | 161 (87.03) | 81.31 | 91.51 |
| **\*Flu-Like Symptoms**<br>**(n=185)** |  |  |  |  |
|  | Experienced flu-like symptoms | 72 (38.92) | 31.85 | 46.35 |
|  | Did not experienced flu-like symptoms | 113 (61.08) | 53.65 | 68.15 |

NB: Variables with (\*) were for the period of six (6) months (January 2020 to June 2020) before the study was conducted.

### SARS CoV-2 Seroprevalence, COVID-19 Asymptomatic Rate and Infection Risks in Niger State

SARS CoV-2 seroprevalence in Niger State was found to be 25.41% (47/185) and 2.16% (4/185) for the positive IgG and IgM respectively as of 26^th^ June 2020- 30^th^ June 2020 (Table 2). The number of participants that did not experienced flu-like symptoms in the last 6 months (January 2020 – June 2020) and have tested positive for SARS-CoV-2 IgG amounted to the COVID-19 complete asymptomatic rate in Niger State (Table 2). The COVID-19 asymptomatic rate in the state was found to be 46.81% (22/47) (Table 2). SARS CoV-2 seroprevalence among age groups, gender and by occupation varied widely (Table 2). Among age groups, the SARS CoV-2 seroprevalence was found to be in order of 30-41 years (33.33%) > 42-53 years (32.42%) > 54-65 years (30%) > 66 years and above (25%) > 6-17 years (19.20%) > 18-29 years (17.65%) > 5 years and below (6.66%) (Table 2). A seroprevalence of 27.18% was recorded for males and 23.17% for females in the state (Table 2). SARS CoV-2 seroprevalence of 37.21% was recorded for health care workers in Niger State (Table 2). Among the non-health care workers in the state, the SARS CoV-2 seroprevalence recorded was 19.01% (Table 2). SARS CoV-2 seroprevalence among the urban dwellers in the state stood at 27.82% while for the rural dwellers, it was 21.42% (Table 2). The same SARS CoV-2 seroprevalence (about 25%) was recorded among the overseas returnees and those that did not travel (Table 2). However, a higher SAR CoV-2 seroprevalence (41.68%) was recorded for those that had contact with the overseas returnees compared to those that did not have contact with the returnees (22.98%) (Table 2).

**Table 2:**
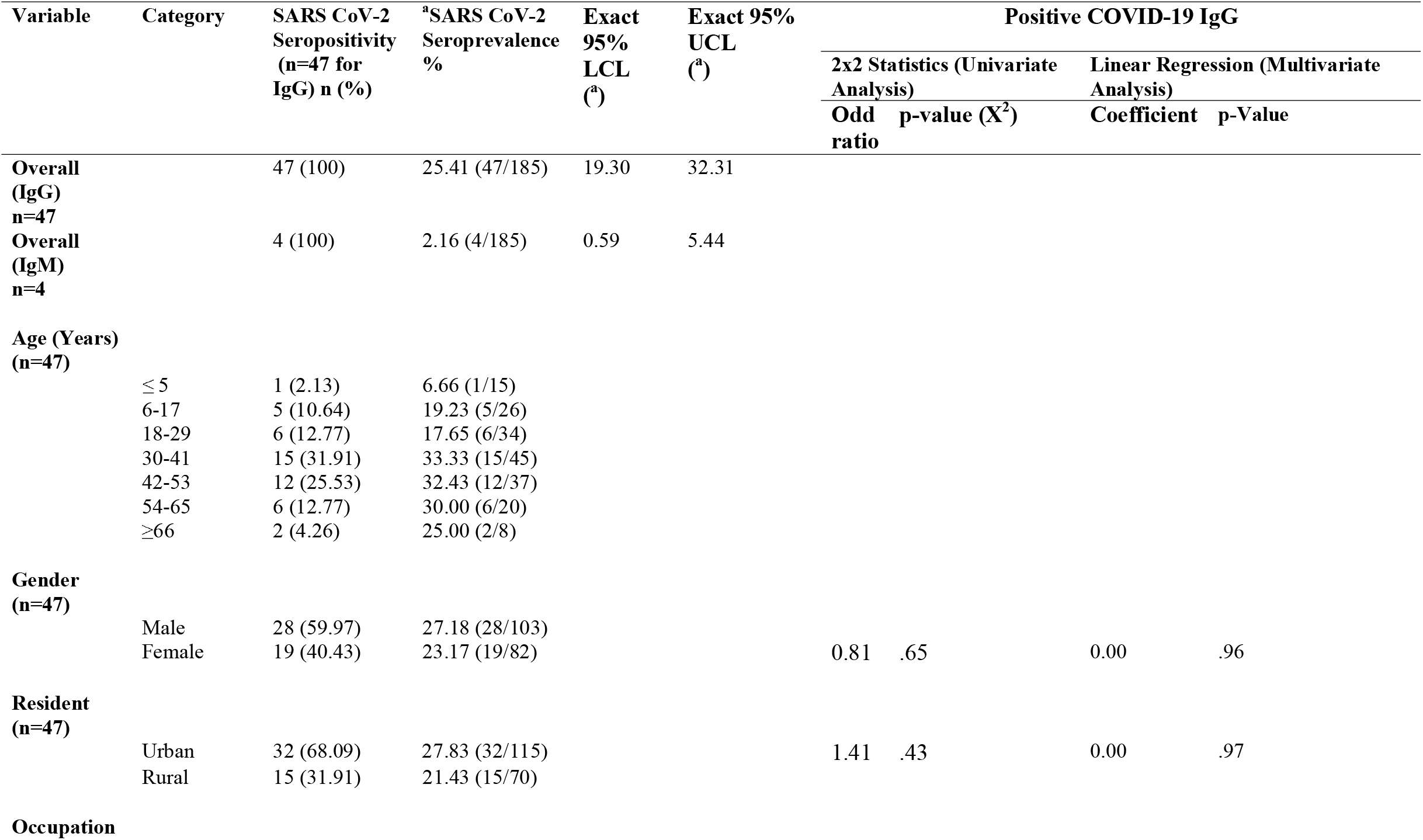
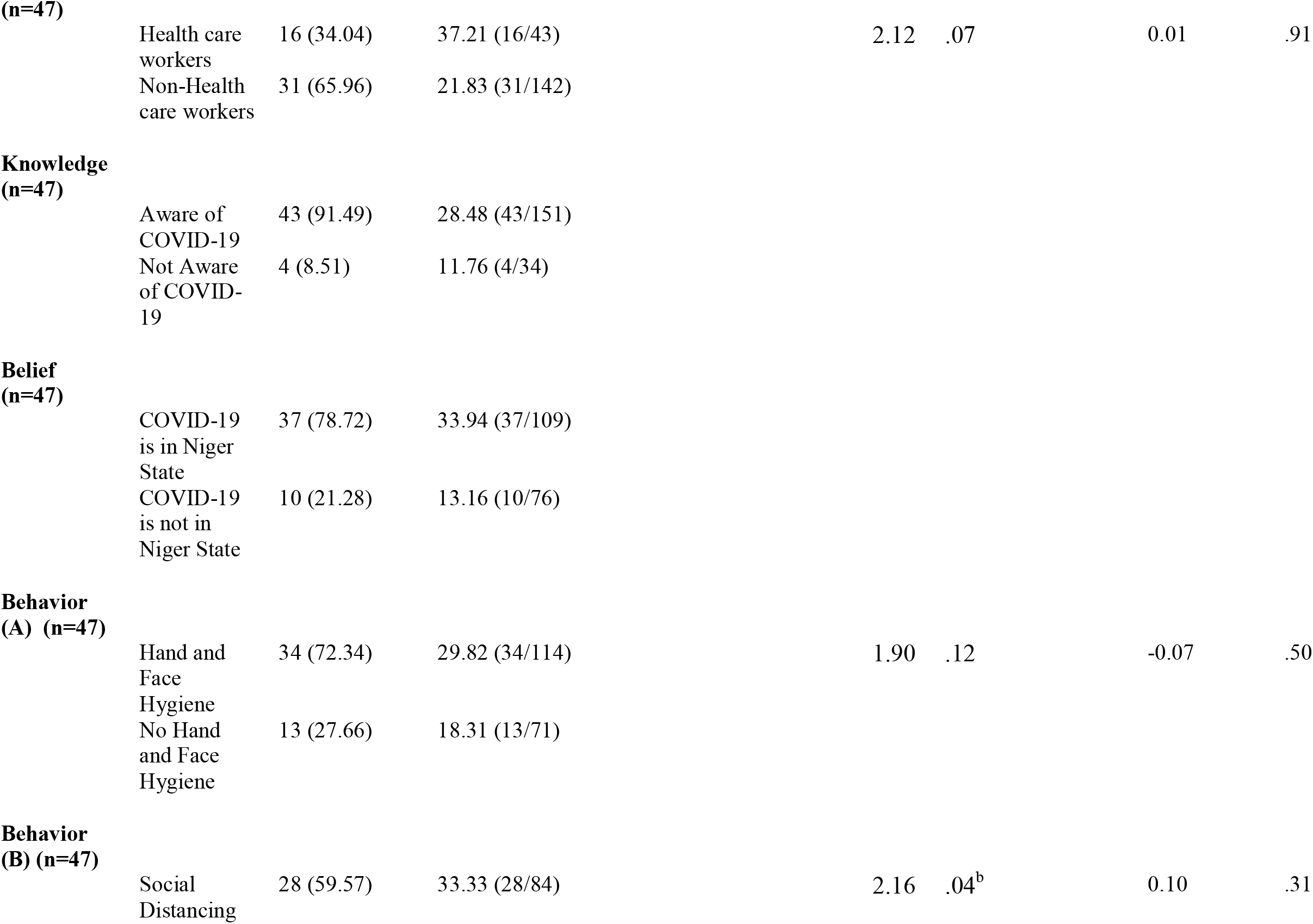
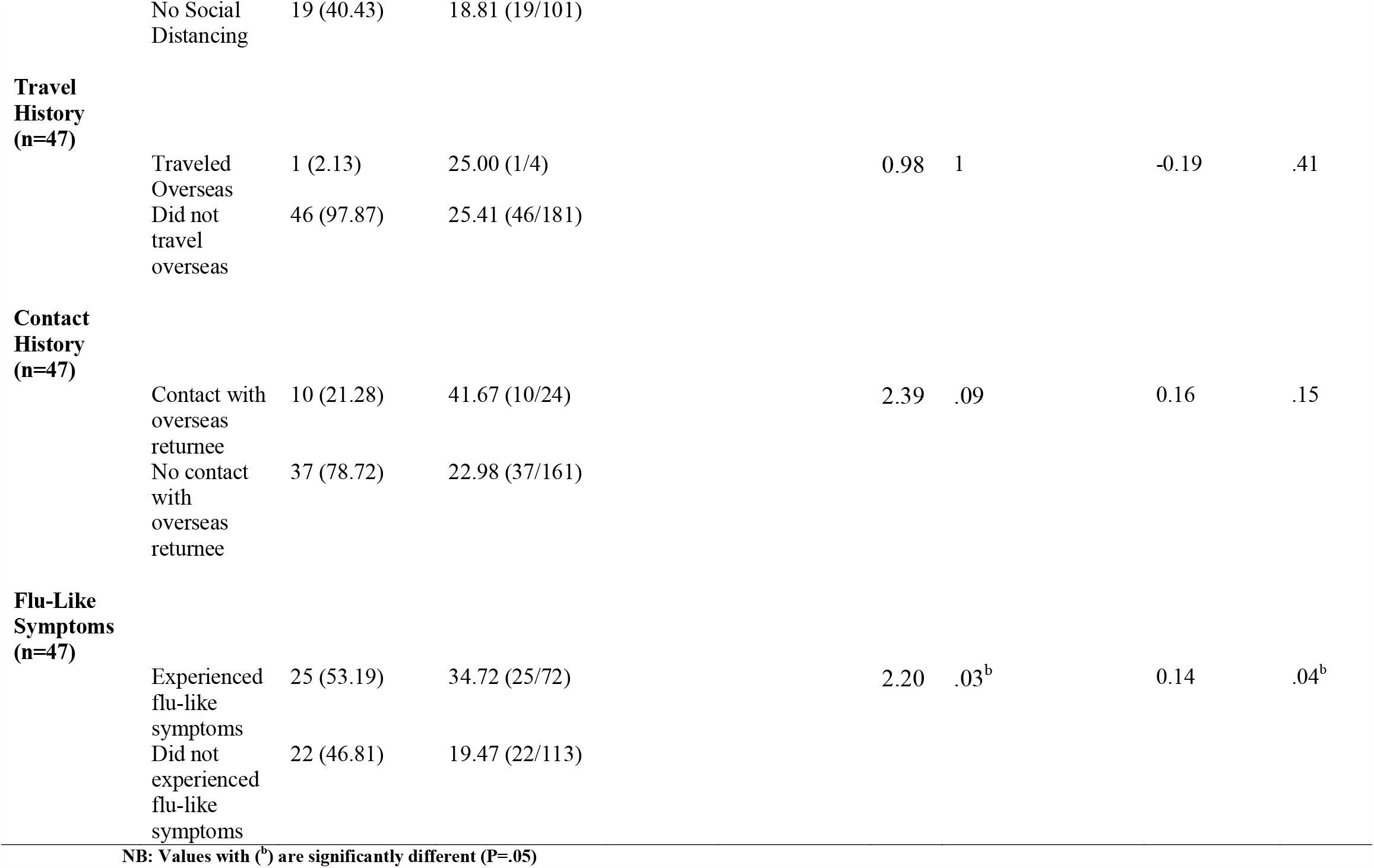
SARS CoV-2 Seroprevalence, Infection Risks and COVID-19 Asymptomatic Rate as of 26^th^ – 30^th^ June 2020 in Niger State, Nigeria

In order to determine the risk factors of SARS CoV-2 infection and also the effectiveness of COVID-19 preventive measures enforced in the state, 2×2 table statistics was used to calculate odd ratios for many scenarios (Table 2). When gender of the participants and Positive COVID-19 IgG results were cross tabulated, the risk ratio recorded for female participants was 0.85 (Table 2).

The risk analyses showed that the chances of infection are almost the same for both urban and rural dwellers in the state even though COVID-19 Seroprevalence among urban dwellers was a little higher than that of rural dwellers (Table 2). Health care workers, those who experienced flu-like symptoms and those that had contact with person (s) that travelled out of Nigeria in the last six (6) months (January 2020 to June 2020) are twice (2 times) at risk of being infected with the virus (Table 2). However, in linear regression multivariate analysis, only “experienced flu-like symptoms” among them was significant at 95 Confidence Interval (Table 2). The risk analyses showed that returning from the overseas did not conferred a protection or posed any increased risk of contracting the virus (Table 2).

## Discussion

### Key findings

SARS CoV-2 seroprevalence pilot study was carried out to know how far the virus has spread in Niger State, Nigeria. Also, to determine patterns, dynamics, and risk factors for COVID-19 in the state. We used a cross sectional study design and Clustered-Stratified-Random sampling strategy to select 185 study participants across three (3) geopolitical zones of the state; this was an effort to have a fair representation of the state even though the sample size was small.

The life expectancy in Nigeria is currently at 55.80 years [8].The gender of the participants reflected the ratio of male to female in Nigeria which is 50.6% male to 49.4% female currently (Table 1) [7]. In Nigeria, currently, 52.0 % of the population lives in urban areas while 48% are in the rural areas [8].

Before COVID-19 vaccines became available, other ways of preventing the transmission of the SARS-CoV-2 (the causative agent of COVID-19 Pandemic) among human populations are social/physical distancing measures and good sanitation and hygiene practices. Adherence to these COVID-19 preventive measures should be impacted by the knowledge and beliefs of people about the disease since the measures involve some behavioural changes and practice. People can only believe what they know (aware of) and can practice when they believe.

There are many reasons why many people did not observe social distancing (Table 1). The first is poverty. The level of poverty in the society is high and many people would have to go out on daily basis to work in order to eat and for their families. Markets are usually open spaces bustling with large crowds of people and where most transactions are done with the physical exchange of cash. This causes situations that defeat the social distancing itself. Also, poverty made many people to turnout in places where food and money are distributed and where physical distancing and other required COVID-19 control measures may not be observed or enforced [17].

The second reason is the prevalence of disbeliefs, myths and conspiracy theories. Many people did not believe in the existence of COVID-19 (Table 1) and especially about the highly contagious nature of the disease. This may be the chief reason why many people did not care to observe social/physical distancing (Table 1) even when not enforced on them at the ATMs, markets, religious gatherings, motor parks, shops, supermarkets etc. Also, myths and conspiracy theories such as COVID-19 does not affect black people, that high environmental temperature and weather will kill off the virus, or that COVID-19 is for rich people and elites are some of the reasons why people are slow in accepting the enormity of the pandemic and therefore usually takes observance of social and physical distancing lightly [17].

Thirdly, no efficient and robust housing and biometric data management systems where everyone is accounted for, especially for the purposes of employment, health, security and social welfare. If these are available, foods and other goods purchased online can be sent to houses with ease. In addition, utilities such as power, water, internet etc, are in most cases not provided or grossly inadequately supplied. It is difficult for people to stay back at home and observe social/physical distancing in such situations [17].

Participants in this study were asked whether they travelled out or have had contact with someone that travelled out of Nigeria since when the pandemic started (last 6 months January-June 2020). The first confirmed case (index case) of COVID-19 in Nigeria was announced on 27^th^ February 2020 when a foreigner in Lagos tested positive for SARS-CoV-2. Soon after that, many people including the contacts of the index case and those that came back into the country and their contacts tested positive for the virus. Although overseas travel prior to the border closures and lockdowns in Nigeria has been associated with the increased chance/risk of contracting COVID-19, this might have changed overtime to more of community transmission of the virus (Table 2).

Looking at the dynamics and trajectory of COVID-19 in Nigeria, in the early days of the pandemic when COVID-19 cases were reported already in urban areas in Nigeria, it is supposed to take few weeks before the virus get to the rural areas [5]. And since the preventive measures such as social/physical distancing (lockdown) and use of face masks were enforced in these early days in most states of Nigeria including the Niger State, living in the rural areas of Niger state and other states of Nigeria ought to have been a protective factor against COVID-19 if the preventive measures were strictly observed. More than half (54.59%) of the participants in this study did not practiced social distancing (Table 1) at any time since the pandemic started even as the lockdown was once enforced in the state; this may be the reason why the risk of infection with virus is the same for the urban and rural dwellers who may be less observant of the preventive measures (Table 2). It is also an indication of community spread of the SARS-CoV-2 in Niger State.

Also, the participants were asked whether they have had flu-like experience in the last 6 months (January 2020 to June 2020) since when the COVID-19 index case was announced in Nigeria; this helped to deduce the rate of COVID-19 asymptomatic rate in Niger State which was 47% (Table 2). Other SARS CoV-2 sero-surveys across the world reported similar high asymptomatic rate of COVID-19 [18-20]. It has been reported that majority of infected people with SARS-CoV-2 (about 50-75%) are usually asymptomatic [21,22].

The seroprevalence of SARS-CoV-2 in Niger State was found to be 25.41% and 2.16% for the positive IgG and IgM respectively. The observed seroprevalence is higher than most of the SARS CoV-2 sero-surveys carried out almost at the same time in other parts of the world [19,20,23] and only a study in India reported a higher seroprevalence of 54.1% [24]. However, by this 25.41% SARS CoV-2 seroprevalence observed in Niger state and also considering the COVID-19 reported cases for Niger State and Nigeria as of 30^th^ June 2020 (when this study was conducted) and 21^st^ December 2020, it means SARS CoV-2 infection occurred very far more than the reported cases in the State and Nigeria [5]. Our data suggest that >5000 times more SARS-CoV-2 infections occurred than the number of reported cases in Niger State or >900 times more SARS-CoV-2 infections occurred than the number of reported cases in Nigeria. The high SARS CoV-2 seroprevalence observed in this study may be due to many factors including high COVID-19 asymptomatic rate and high lack of observance of social distancing in the State as observed also in this study. Unknown high proportion of asymptomatic COVI-19 cases may not be diagnosed and not reported and therefore, our observed SARS CoV-2 seroprevalence in the State will be more reliable and closer to the actual true prevalence of the disease than the official reported cases. As of 21^st^ December 2020, and based on the COVID-19 reported cases and deaths, the fatality rates for COVID-19 in Niger state and Nigeria stood at 3.15% and 1.54% respectively [5]. However, when the observed 25.41% SARS CoV-2 seroprevalence was considered the fatality rates drastically reduced to 0.0009% and 0.024% for Niger state and Nigeria, respectively.

Usually, after 2-3 days of the onset of symptoms of COVID-19 or after 10 days in case of asymptomatic COVID-19, IgM become detectable in whole blood/serum/plasma of the patients [11,16]. The IgM level in the blood peaked after 14 days of the infection with the SARS-CoV-2 and it disappears after 28 days of the infection [11,16]. However, IgG production starts after 14 days of infection and it remains in the blood for long time immunity [11,16]. The timeline for production and disappearance of IgG and IgM are useful in the interpretation of the COVID-19 IgG and IgM Rapid Test. The test kit detecting only IgM means that the participant/patient is at the early stage of the infection while the kit detecting only IgM means that participant/patient have had a past infection and have recovered. However, the test kits detecting both the IgG and IgM at the same time means the participant/patient may be in the recovery stage of the infection. In this study, IgG only and IgG plus IgM were observed. This means overwhelming majority of the participants that tested positive (positive IgG only) for the tests have had past infections and have recovered (Table 2). And only very few (positive for both IgG and IgM) were just recovering from the infection (Table 2).

The SARS CoV-2 seroprevalence among the age groups and gender correlated with the most mobile/active of the age groups and gender in our society (Table 2). Age groups 30-41 years, 42-53 years and 54-65 years are the most mobile of the age groups while men are more mobile than the women and therefore can contract the virus easily. We observed less likely odds of contracting COVID-19 among females compared to the males (Table 2). This means that being a female is a protective factor against the infection of SARS-CoV-2 in Niger State. This also correlated with the COVID-19 Seroprevalence recorded among males and females participants (Table 2). The less risk of infection by the females in this study maybe due to the physical attributes such as the less mobility and activity of the females compared to that of males in our society. Generally, around the world, the case fatality of COVID-19 varied widely (1-20%) with more cases and fatalities observed in males compared to females [25,26].

High SARS CoV-2 seroprevalence (37%) and double odds of contracting COVID-19 among health care workers (Table 2) were observed. It is expected for the health care workers to have higher COVID-19 prevalence compared to the non-health care workers because they are the frontline workers responsible for the diagnosis, treatment, and management of patients including symptomatic and asymptomatic COVID-19 patients [27-29]. These enormous essential tasks for controlling the COVID-19 Pandemic coupled with the inadequate or lack of PPE in some instances and high asymptomatic rate of COVID-19 among people put the health care workers at greater risk of contracting and transmitting the disease. Double odds of being positive for SARS CoV-2 were also observed for the participants that experienced flu-like symptoms and did social distancing since when pandemic started (January 2020 to June 2020) (Table 2). The double odd for the flu-like symptoms was expected and in line with our findings that about 50% of the SARS CoV-2 infections in the state were asymptomatic(Table 2). However, the double odd for the social distancing is not correct, maybe due to confounding issues; this was confirmed in linear regression multivariate analysis Table 2.

### Limitations

The study has some limitations. Firstly, selection bias may exist as it was more difficult to recruit participants that were 5 years and below. Secondly, the sample size used in this pilot study and number SARS CoV-2 Rapid Test Kits used for validation are small. The SARS CoV-2 Rapid Tests Kits that are suitable for epidemiological studies are highly costly; this limits the sample size of this pilot studies. Thirdly, to get a quick understanding of the levels of knowledge, attitude and practice about COVID-19, we kept the questionnaire short and simple, which might have limited the depth of the study.

## Conclusions

To the best of our knowledge, this study is the first pilot SARS CoV-2 seroprevalence data for Nigeria. The study revealed SARS CoV-2 seroprevalence, patterns, dynamics, and risk factors in Niger state, Nigeria. The seroprevalence of SARS CoV-2 in Niger State was found to be 25.41% and 2.16% for the positive IgG and IgM respectively. Seroprevalence among age groups, gender and by occupation varied widely due to the differences in mobility and activity as well as the occupational exposures and hazards. The COVID-19 asymptomatic rate in the state was found to be 46.81%. The risk analyses showed that the chances of infection are almost the same for both urban and rural dwellers in the state. However, health care workers, those that experienced flu-like symptoms and those that had contact with person (s) that travelled out of Nigeria in the last six (6) months are twice (2 times) at risk of being infected with the virus. More than half (54.59%) of the participants in this study did not practiced social distancing at any time since the pandemic started. The observed Niger State SARS-CoV-2 seroprevalence and infection patterns means that the virus is widely spread, more SARS CoV-2 infections occurred than the reported cases and high asymptomatic COVID-19 across the state. Our data suggest that >5000 times more SARS-CoV-2 infections occurred than the number of reported cases in Niger State or >900 times more SARS-CoV-2 infections occurred than the number of reported cases in Nigeria.

## Data Availability

All data referred to in the manuscript are available

## Acknowledgement

This work was funded by the Ibrahim Badamasi Babangida University, Lapai, Niger State, Nigeria. The research team would like to thank the Niger State Ministry of Health for their support throughout the study.

